# Developing a Data Note reporting guideline for qualitative health and social care research datasets (the DeNOTE study): A study protocol

**DOI:** 10.1101/2025.02.04.25321640

**Authors:** Hannah A. Long, Elaine Toomey, Fiona Stevenson, Joanna M. Brooks, Andrew J. Stewart, David P. French

**Affiliations:** Healthier Futures Research Platform, Division of Nursing, Midwifery, and Social Work, School of Health Sciences, Faculty of Biology, Medicine and Health, University of Manchester, UK; Centre for Health Research Methodology, School of Nursing, and Midwifery, University of Galway, Ireland; Department of Primary Care and Population Health, University College London, UK; Manchester Centre for Health Psychology, Division of Psychology and Mental Health, School of Health Sciences, Faculty of Biology, Medicine and Health, University of Manchester, UK; Department of Computer Science, School of Engineering, Faculty of Science and Engineering, University of Manchester, UK

**Keywords:** qualitative research, health and social care, open data, open research, reporting guideline, data note, data article, data paper, DeNOTE

## Abstract

**Background:** Data Note articles provide concise descriptions of research datasets, detailing how and why the data were created and facilitating reuse of the data. Additionally, Data Notes are typically believed to have the added value of increasing research transparency and making the data comply with the FAIR principles (i.e., Findable, Accessible, Interoperable, and Reusable). However, existing guidelines and templates for Data Note articles are designed for quantitative research datasets and there are no equivalents for qualitative research datasets.

**Aim:** To develop a novel reporting guideline for Data Note articles describing qualitative health and social care datasets (i.e., the DeNOTE reporting guideline).

**Methods:** Our plans to develop the DeNOTE reporting guideline have been registered on the EQUATOR (Enhancing the QUAlity and Transparency Of health Research) Network. In summary, the DeNOTE study consists of (a) a rapid scoping exercise of existing literature, resources, and expert knowledge to identify and synthesise ‘items’ or ‘statements’ of a reporting guideline that are relevant to a Data Note article describing qualitative health and social care data, (b) an online questionnaire with expert participants to rate their agreement with items identified in (a) and to propose new or amended items, (c) an online workshop with participants to further co-develop the reporting items, with the aim to reach consensus on the items, (d) eliciting feedback from participants on the draft reporting guideline, and (e) finalising the DeNOTE reporting guideline.

**Conclusion:** Our goal is to produce a novel reporting guideline to support researchers when producing a Data Note article describing qualitative health and social care research data. In the process, we will produce tailored open research infrastructure to better support qualitative researchers engaging in open research practices.

## BACKGROUND

The open research movement is an ongoing transdisciplinary effort to make research processes and outcomes more transparent and accessible to other academics, policymakers, professionals, and the public(1). In quantitative research communities, recommended best practice is data sharing, whereby researchers make their data collection instruments, datasets, and analyses available in public repositories. Open data practices are seen as important for advancing scientific knowledge, fostering collaboration and innovation, and ensuring transparency and accountability in research. Prominent organisations and funding bodies - such as UK Research and Innovation (UKRI), Economic and Social Research Council (ESRC), and the National Institute for Health and Care Research (NIHR) in the UK, as well as the National Institutes of Health (NIH) in the USA - and leading publishers - such as Elsevier, Springer, and Wiley - have endorsed the FAIR (Findable, Accessible, Interoperable, Reusable) principles (2) and embraced policies that advocate for or mandate open data.

Although open research practices initially originated in response to concerns regarding quantitative research, the core tenets of open research of promoting transparency, accessibility, rigour, and collaboration are of relevance to all research(3). However, current open data requirements often overlook key epistemological and methodological differences between quantitative and qualitative research. The expectation that full qualitative datasets should be made available alongside research publications raises legitimate sensitivities and ethical concerns (3–5). Even with thorough de-identification, the potential for identifying participants in qualitative studies remains high, given the detailed and deeply personal nature of the data, with additional issues for certain data formats such as video-recordings of participants (6,7). Further, the prospect of sharing qualitative data for replication purposes is in conceptual conflict with the core principles of most qualitative methodologies, which emphasise the researcher’s unique interpretative role throughout study design, data collection, and analysis (4,8). While open data practices are advocated for in the name of transparency, the question remains whether sharing complete qualitative datasets is necessary or appropriate for ensuring transparency in qualitative research (4). Despite the ongoing discussion and lack of consensus on open qualitative data sharing, there are nevertheless several clear benefits to sharing qualitative data. These include maximising the impact and cost-efficiency of data collection, facilitating secondary analyses and synthesis reviews, offering students and trainees real-world datasets for developing analytic skills, and increasing public trust in science through (the appearance of) greater transparency (4,9,10). Research participants agree to qualitative data sharing, motivated by the prospect of helping others and improving research(11). As researchers, we have a responsibility to maximise the time and effort participants give to research, while protecting their privacy(3). Robust guidance for those who do wish to share their data is thus needed.

As the open research movement advances, an increasing number of resources and research infrastructures have been established to support open data sharing. The issue is that many of these resources have roots in quantitative paradigms, favour quantitative research methods, and are thus generally unsuited to qualitative research. One example is the current guidance on Data Notes (also known as Data Papers, Data Articles, or Data Descriptors). Data Notes (DNs) are peer-reviewed articles that succinctly describe how and why an archived research dataset was created, with the purpose of enhancing research transparency, promoting data reuse, and making the data FAIR (12). Organisations and academic publishers are increasingly advocating for DNs (e.g., (13–15)), while certain journals have mandated their submission alongside research publications (e.g., (16)) and have requested DNs for special issues (e.g., (12,17)).

Existing guidance to authors producing DNs ranges from written instructions on the information to include, to explicit and prescriptive DN templates (15,18,19). These resources incorporate information and elements (e.g., missing data handling) exclusive to quantitative research datasets. Other elements (e.g., data validation) do not align with qualitative conventions or account for processes fundamental to qualitative research (e.g., protecting participant identity). While some aspects of these guidelines and templates (e.g., rationale for dataset creation) could apply to qualitative data, the lack of alignment with qualitative methods makes it unclear how qualitative researchers can adapt these guidelines to suit the nature of their research data. Rather than imposing ill-suited templates on qualitative researchers, there is a clear need for a novel counterpart: a DN reporting guideline specifically designed for qualitative health and social care data. To the best of our knowledge, no such guideline exists. This research is timely as a new repository is currently being developed specifically for qualitative health and social care data storage (20).

It follows that we support the British Psychological Society’s 2020 call for tailored open research infrastructure to better support qualitative researchers (21). A DN reporting guideline for qualitative datasets needs to reflect the distinct characteristics, intricacies, and methodological practices of qualitative research. Therefore, this study will endeavour to identify the important items of a reporting guideline for DN articles describing qualitative health and social care datasets. We have chosen a focus on health and social care research data on the grounds of the unique legal and ethical complexities and sensitivities of such data. We wish to adopt a definition of guidelines as ‘recommendations’ (not rules) that offer a flexible approach to reporting research, emphasising *process* overcome *outcome* (22). Unlike criteria or standards, which are more fixed and measurable, guidelines encourage interpretative judgements and expert discretion. They foster a more nuanced and thoughtful approach, often relying on the expertise and experience of authors, and are not intended to be strictly prescriptive or enforcing thoughtless adherence(22). To achieve our goal, the study will engage directly with key actors who are likely to produce and use DN articles (e.g., qualitative health and social care researchers, open research advocates) or who have their data represented in DN articles (e.g., patients and the public). This will ensure the DN reporting guideline is relevant, comprehensive, and easy to understand for the qualitative health and social care research community.

### Aim

The aim of the present study is to produce a novel reporting guideline for DN articles describing qualitative health and social care research datasets. The specific objectives are to:

a. Conduct a rapid scoping exercise to identify existing DN reporting guidelines, templates, and guides for authors in health, social care, and allied social science disciplines.
b. Extract the writing guidance and reporting items from existing literature identified in (a), assess its relevance and suitability for reporting qualitative health and social care research datasets, and synthesise these data into a list of potential reporting items.
c. Use questionnaire and group consensus methods with expert participants to review and consolidate the results of (b), to identify further reporting items, and aim to reach consensus on the essential items for a DN reporting guideline for qualitative health and social care research datasets.
d. Produce a draft DN reporting guideline and seek feedback from expert participants.
e. Produce a finalised, exemplar reporting guideline for DNs describing qualitative health and social care research datasets, with endorsement from expert participants.

## METHODS

### Study design

The present study will use mixed methods in a sequential design. The study will follow the EQUATOR (Enhancing the QUAlity and Transparency Of health Research) Network methodology for developing article reporting guidelines as far as possible (23). Broadly, this involves (i) identifying the need for a reporting guideline, (ii) ‘preparing the ground’ and identifying suitable experts, (iii) developing the reporting guideline, and (iii) writing up and publishing the reporting guideline. In accordance with the EQUATOR approach, our intention to develop the DeNOTE reporting guideline is registered with the EQUATOR Network. This is to raise awareness of the novel guideline under development, to prevent any duplication, and to ensure transparency about the process.

### Procedure

The specific steps to develop an article reporting guideline for health and social care research are summarised in *Table 1* (23). The four stages of the study procedure are described below.

**Table 1.**
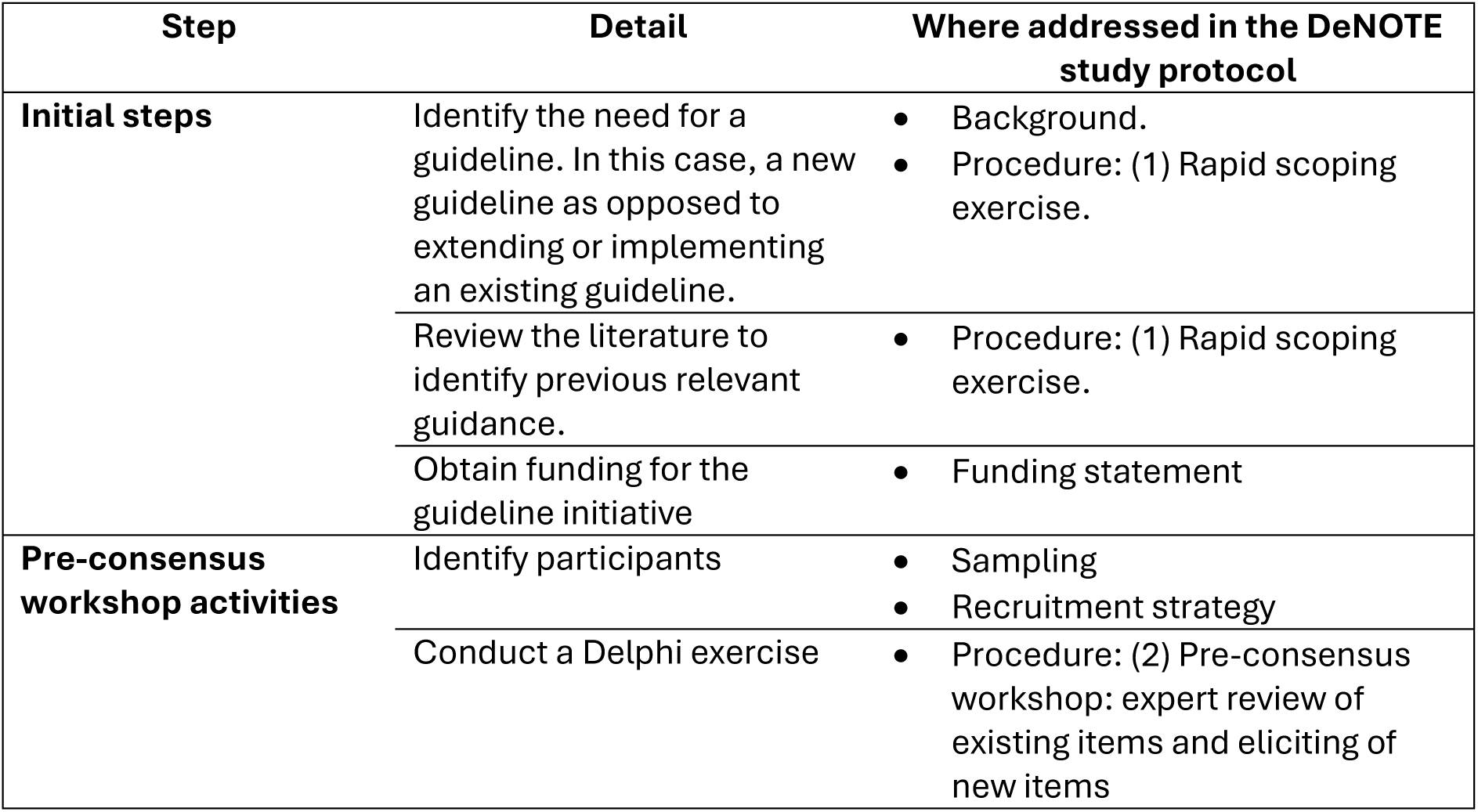
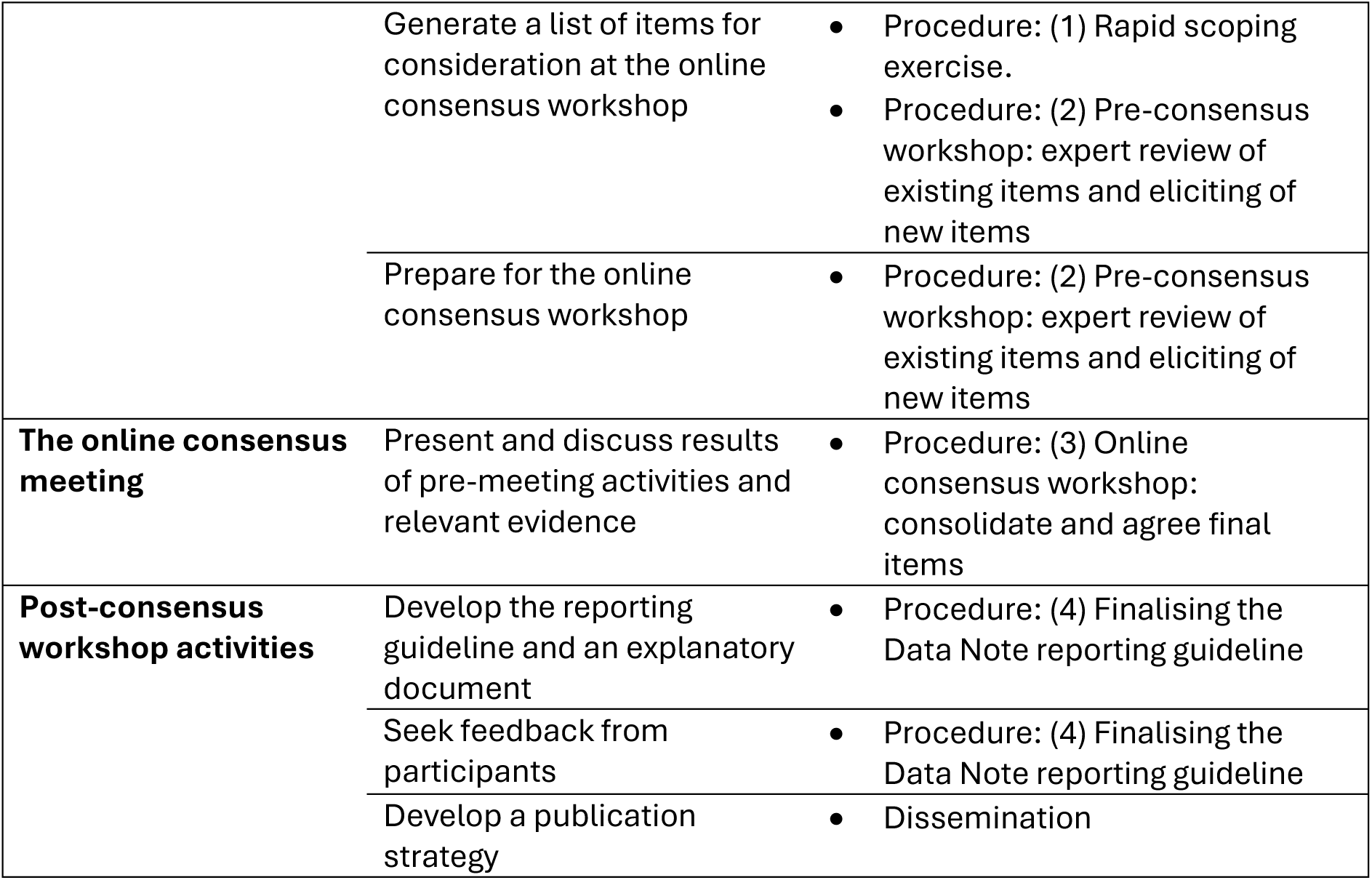
Summary of recommended steps for developing a health research reporting guideline (adapted from Moher et al., 2010).

#### 1) Rapid scoping exercise to identify existing relevant reporting guidelines and guidance

To the best of our knowledge, there are no existing reporting guidelines (or other formal guidance or templates) for DN articles specific to qualitative health and social care research datasets. To investigate this further, we will conduct a rapid scoping exercise to identify relevant documents and publications on the reporting of DN articles. This is necessary to describe what guidance is currently in place, the extent to which this guidance is relevant and applicable to describing qualitative health and social care data and, where appropriate, to use this guidance to inform the next stages of the study.

The steps of the rapid scoping exercise will be fourfold. Firstly, Scopus will be searched to identify existing DN-specific reporting guidelines, guidance to authors, article templates, or similar. The search strategy will include appropriate thesaurus terms, free text terms, and key words for DNs (e.g., “data note”, “data paper”, “data article” and “data descriptor”) and guides for authors (e.g., “guide*”, “reporting criteri*”, “template*”, “guidance” and “guideline*”) (Appendix A). The searches will be limited to documents published in English. There will be no restriction on publication type. To facilitate rapidity, up to 1000 search results will be retrieved and screened. If the searches return many more results than 1000, other limits may be applied (e.g., a date/year restriction).

Secondly, the EQUATOR Network database of journal article reporting guidelines will be searched to identify relevant reporting guidelines for primary qualitative research. Three relevant reporting guidelines are already known to the DeNOTE steering committee: (1) the American Psychological Association (APA) style Journal Article Reporting Standards (JARS) (24), (2) the COnsolidated criteria for REporting Qualitative research (COREQ) (25), and (3) the Reflexive Thematic Analysis Reporting Guidelines (RTARG) (26). It is possible that other relevant reporting guidelines exist. It is believed that these guidelines are likely to include existing items or criteria related to the reporting of data or other study features that are transferrable or extendable to a reporting guideline for DNs of qualitative datasets.

Thirdly, the websites of relevant publishers and journals will be hand searched for suitable DN reporting guidelines and/or guidance to authors producing DNs. Target publishers and journals will include, but may not be limited to, the highest ranking (based on impact factor) health, social care, and social science outlets, and the top Open Access publishers in these fields (e.g., those endorsing the Transparency and Openness Promotion (TOP) Guidelines, developed by a community working group in conjunction with the Centre for Open Science (1)). Publishers will be contacted to inquire whether they provide DN writing guidance to authors. Further, the websites of leading relevant research funders, organisations, and bodies (e.g., NIHR Open Research, Open Research EU), will be searched to identify relevant guidance to authors reporting DN articles (including that which is published in webpage form only).

Finally, relevant experts known to the DeNOTE steering committee will be contacted to recommend existing DN article reporting guidelines and/or guidance documents to authors producing DNs for qualitative health and social care datasets. These experts may include, but not be limited to, individual academics known to be actively publishing qualitative health and social care research and open research, as well as members of relevant special interest, committee, and society groups.

All identified documents will be imported into appropriate reference management software (e.g., EndNote and/or Rayyan). One member of the DeNOTE steering committee will screen all documents against prespecified eligibility criteria. In instances where document eligibility is unclear, the document will be discussed by two or more steering committee members until a decision is agreed. The eligibility criteria for the inclusion of documents are:

a. Documents must explicitly present guidelines (including criteria, items, domains, flow diagrams, templates, or instructive statements) for the reporting of DN articles. The DN guidance in question must relate to data generated in human health, social care, and/or social sciences research, *or* be general and generic (i.e., subject- or discipline-free). Documents that do not explicitly address the *reporting* of data will be excluded (e.g., those providing general guidance or policies on open data, data sharing, and data management).
b. The guidelines can pertain to either quantitative or qualitative research data.
c. Existing reporting guidelines (i.e., those registered in the EQUATOR Network database) for primary qualitative research studies, covering all or part of the area in question, are eligible. This excludes reporting guidelines for systematic reviews of qualitative research (also known as qualitative evidence syntheses and qualitative meta syntheses), as well as other review types (e.g., rapid, scoping, and umbrella), and qualitative study protocols.
d. The document must be written in English.
e. A full text version or equivalent must be available.

All identified documents will be reviewed for relevant information and synthesised. A systematic content analysis of the eligible documents will be performed to identify and extract domains, items (or similar), and accompanying item statements. A domain may refer to a broad category or area of focus that encompasses multiple related aspects (items) (e.g., ‘data collection’), whereas items represent specific discrete elements or details that may or may not fall under a domain (e.g., ‘describe the sampling method used to recruit participants’). The domains and items from each document will be assessed in terms of whether they are relevant, partially relevant, or of no/unclear relevance (and thus not transferrable) to the reporting of qualitative health and social care data. We will endeavour to determine whether existing guidance involved patient and public contributors in its development. It is anticipated that DN guidance pertaining to data generated in human health, social care, and social sciences research will be assessed first, as this will be the most relevant and transferrable. A preliminary item list will be produced. This domain/item list will then be compared and contrasted with general DN guidance to identify any additional relevant domains/items. It is anticipated that there will be a degree of repetition in the content of eligible documents. To manage this, a pragmatic approach will be taken; the DeNOTE steering committee may, for example, merge similar domains or items, separating those that cover multiple distinct aspects of research practice, and selecting the best representation of any duplicated domains or items found in the literature. The resulting domain and item list (and any accompanying item statements) will form the basis for the next stage of the study.

#### 2) Pre-consensus workshop: expert review of existing items and eliciting of new items

Expert participants will be identified and purposively recruited to the study. The first task for participants involves completing an online questionnaire. Use of an online questionnaire allows input from international experts without geographical constraints.

The questionnaire will be piloted with a small group of academics known to the DeNOTE steering committee and amended where necessary before being shared with participants.

Participants will receive an email with a link to an online questionnaire (hosted on Qualtrics, Microsoft Forms, or similar) and other materials relevant to the study and research task. Participants will receive one reminder email at approximately two weeks if no reply has been received.

In the questionnaire, participants will be asked to review, rate, and indicate items needed for a DN reporting guideline for qualitative health and social care data. The questionnaire will present the list of reporting items identified from existing literature during the scoping exercise. Participants will be invited to rate the importance of items and to propose new or amended items. This will allow participants to propose and frame what they consider to be important to the development of the reporting guideline. Participants will be encouraged to include textual item statements that explain the item (e.g., a sentence or two to summarise the essential concept, feature, or indicator in the item). The items should be designed to inform, steer, and support individuals who are reporting a DN article. Views of participants may be further elicited using a small number of open and/or close-ended questions to gain insight into the sorts of issues relevant to the reporting guideline, which may need to be discussed at the online consensus workshop, as well as participants’ experiences of using reporting guidelines and producing DN articles in their research. Therefore, the questionnaire exercise will identify specific items and issues relevant to producing a reporting guideline for DNs of qualitative health and social care research.

It is expected that some items will be rated as less important than others. It is also possible that there will be a degree of repetition in participants’ suggestions for new items. Therefore, depending on the results of the questionnaire, it may be deemed pragmatic for the DeNOTE steering committee to reduce the item list to reach a more manageable number for discussion at the consensus workshop. When many items exist, a classification scheme for selecting items for inclusion is encouraged(23). A report of the questionnaire procedure and the list of proposed items will be shared with participants prior to the consensus workshop, alongside information on the format and logistics of the workshop.

#### 3) Online consensus workshop: consolidate and agree final items

The same participants will be invited to take part in an online consensus workshop held on Microsoft Teams in early 2025(23). Inviting the same participants to both the questionnaire and the workshop ensures continuity and consistency in the exploration of ideas and fosters a greater sense of group ownership over the ideas(27). It allows participants to clarify and refine their initial responses during the workshop. The central objective of the consensus workshop will be to explore and as far as possible combine the views of experts to reach agreement on the precise items that the reporting guideline will cover (i.e., the scope of the items and their guiding statements). This will mainly involve reviewing, consolidating, and reducing the list of potential items identified via the online questionnaire, which will form the central part of the reporting guideline. The optimal format and phrasing of the reporting guideline will also be discussed. The online workshop format will consist of a combination of small group discussions and plenary sessions. The workshop will be transcribed using the inbuilt Microsoft Teams transcription function and be audio- and video-recorded in order to later check the transcription for accuracy.

At the start of the workshop, the DeNOTE steering committee will present the background information to the study, any relevant empirical evidence from the literature, the rationale for including items in a reporting guideline, and the results of the online questionnaire. Participants will have the opportunity to seek clarification on any unclear results.

Participants will then be invited to work together in small groups to discuss, review, and refine the items proposed in the online questionnaire. Depending on the results of the online questionnaire, it may be pragmatic to assign different clusters of related items to different groups of participants (with diverse expertise) to discuss. A key feature will be eliciting from participants what they consider to be the most important items and issues. These discussions will likely focus on the content of item information, rather than the precise wording of the items at this stage. It will be emphasised that, typically, the final items to be included should represent a minimum essential set of items to report (23), and therefore a discussion of participants’ perspectives on this will be highly valuable.

A member of the DeNOTE steering committee will chair each group discussion. If necessary, prompt questions may include: What makes certain items more important than others? Which items do you consider essential? Are the proposed items sufficiently distinct? Are any items too similar? Is anything missing? The suggestions made by the small groups will then be presented back to the whole group and discussed in a plenary session. This will also allow for any within-group disagreements to be considered and resolved as a whole group.

The revised list of items will be presented back to the whole group. Participants will vote privately via an online form towards the end of the workshop to determine which items to include in the final reporting guideline. In the case of mixed consensus, further discussion may be required, either as a whole group or in smaller groups, to reach verbal consensus, and/or further voting (either during the workshop or via a follow-up questionnaire sent after the workshop). If agreement is not reached, the process will nevertheless identify where consensus has not been possible.

#### 4) Finalising the Data Note reporting guideline

Following the consensus workshop, the DeNOTE steering committee will serve as the ‘writing group’ responsible for drafting the proposed DN reporting guideline. Other individuals who can make a clear contribution to the writing of the reporting guideline may be included in the writing group. The process is likely to involve several iterations. The writing group will aim to translate the workshop discussions into clear, precise, and unambiguous reporting items, while also determining the most appropriate order for the items and structure of the guideline.

The draft DN reporting guideline will be circulated to the workshop participants (‘the DeNOTE group’) for a minimum of one round of feedback. Based on this input, the writing group will finalise the DN reporting guideline. All participants will be asked to formally approve the final version via email before it is submitted to a journal for publication. Participants will be given two opportunities (email reminders) to do this; if no response is received, the final version will be submitted without participants’ explicit endorsement.

### Sampling

Developing a robust reporting guideline needs input from a wide range of people with differing areas of expertise and who represent different disciplines, organisations, and countries (23). Approximately 25-30 experts from multidisciplinary backgrounds will be recruited to the study. The aim is to identify key individuals with experience and expertise that reflects the reporting guideline in development. This involves individuals who are likely to be key users, implementers, and/or disseminators of the final reporting guideline, including research participants and those involved in scientific research funding and publishing. This is to ensure that the reporting guideline reflects user needs and is more likely to be adopted into research practices (27). The proportion of content experts (i.e., qualitative health and social care researchers) should be at least 25% of the group (23).

People will be purposively recruited to ensure a diverse sample based on their disciplinary background, experience, and expertise. A sampling framework will be developed to ensure the spread of the following characteristics: (i) career stage (e.g., early, middle, and senior researchers who are likely to be the primary producers of DN articles); (ii) experience with qualitative health, social care, and allied social sciences research or open qualitative research (as researchers or as patient and public contributors taking part in qualitative research); (iii) experience on qualitative health, social care, and social sciences funding body panels; (iv) experience as editors of qualitative health, social care, and social sciences research journals; (v) experience on committees, in special interest groups, or in advisory groups for relevant professional bodies (e.g., European Society for Health Psychology Open Science Special Interest Group; NIHR Open Research Advisory Group); (vi) experience in data management, storage, and stewardship (e.g., university librarians, data stewards) or research ethics and information governance. Patient and public contributors with knowledge of and/or experience in qualitative research will be involved to include the perspectives and experiences of those who participate in or collaborate on qualitative health and social care research. Their involvement is particularly critical in representing the concerns, priorities and expectations of individuals who contribute data to research studies and are therefore directly impacted by data management and storage policies and procedures.

All participants should have contact details in the public domain (e.g., higher education institution email address) or available through contacts known to the DeNOTE steering committee, and be able to read, write and understand English to a sufficient level to participate.

### Recruitment strategy

The recruitment strategy will be threefold to ensure the inclusion of people with a diverse range of relevant experience. Firstly, we will leverage the multidisciplinary knowledge and expertise within the DeNOTE steering committee, utilising our professional networks, to identify suitable participants. These networks, which span various disciplines in health, social care, psychology, sociology, and open research, will provide insights into potential experts who are not only knowledgeable of the breadth of qualitative research methods used in health and social care research, but who are also actively engaged in the current discourse on qualitative and open research. Further, we will leverage our existing networks to identify patient and public contributors with knowledge of and/or experience in qualitative research to allow the experiences of people who take part in research, particularly qualitative research, to be reflected in the final reporting guideline.

Secondly, we will examine authorship of key publications in qualitative health and social care research and open research. Particular attention will be given to authors of articles that examine the intersection of qualitative methods and open research, especially those that highlight the challenges and opportunities this integration presents. This will ensure the inclusion of individuals who are contributing to the advancement of these fields and who can bring nuanced perspectives to the discussion.

Finally, we will use our networks to identify key individuals who manage qualitative health and social care data and data searches (e.g., librarians and data stewards). We will contact university ethics committees and National Health Service Research Ethics Committees to identify individuals with research ethics and information governance expertise. We will examine websites of interest (e.g., prominent research funding bodies in health and social care research, leading academic publishers, and other relevant professional bodies) to identify representatives with an interest in qualitative health and social care data reuse. We will employ snowball sampling techniques to recruit further potential participants via the individuals we invite to the study.

Potential participants will be contacted via email and invited to the study by a member of the DeNOTE steering committee. The invitation email will include the study protocol and participant information sheet. The invitation email will ask experts to read through the study information in their own time and to respond to the invitation by replying to the email. Invitations to the study will also include requests for recommendations of colleagues or contacts who may contribute usefully to the study. Potential participants will not be pursued beyond two reminder emails.

## ETHICAL CONSIDERATIONS

Ethical approval for the study was granted by the Proportionate University of Manchester Research Ethics Committee (reference 2024-21401-38793). Potential participants will receive an email with a study invitation and participant information sheet. The participant information sheet will describe the aims and procedure of the study, to give potential participants enough information to allow them to make an informed decision as to whether to take part. The initial email and study materials will include the contact details (i.e., email address) of the lead researcher (HAL) so that potential participants can contact her with any questions and/or to express an interest in taking part in the study.

Participants will be asked to provide their informed consent on two occasions: at the start of the online questionnaire and prior to attending the online consensus workshop. Consent procedures will be built into the online questionnaire hosted on Qualtrics/Microsoft Forms, such that participants can only proceed to the questionnaire once their electronic consent has been provided. A consent form (Microsoft Word document) will be emailed to participants before the online consensus workshop, to be completed and returned by email prior to attending the workshop.

Participants will be explicitly informed in the participant information sheet about how their data will be used in the DeNOTE study. Participants will be offered the opportunity of group authorship of the study outputs. Participants who consent to group authorship will be named and listed as part of the DeNOTE group on all study outputs, which include a peer-reviewed journal article of the DN reporting guideline, archived study data, and any other study outputs (e.g., conference proceedings). Participants who do not consent to this role will remain anonymous and confidential in all study outputs. However, the identities of all participants will necessarily be known to those present at the online workshop. This offering is in line with the CRediT authorship taxonomy; participants who consent will classified as an ‘investigator’ for their collective role in constructing the data through their participation (i.e., ‘performing data collection’) (28). This classification includes group authorship of the study outputs in acknowledgement of participants’ contributions.

It is important to note that ‘the DeNOTE group’ is distinct from the writing group. The latter will be responsible for preparing the study outputs (for which participants have the option to claim group authorship) and making final decisions. The opportunity for group authorship will be introduced in the study invitation and further explained in the participant information sheet. Participants will be explicitly informed in the participant information sheet of the conditions that must be met to qualify for group authorship, which include completing all research activities (questionnaire, workshop, and providing feedback on or approving the draft DN reporting guideline).

## RISKS TO PARTICIPANTS

We do not anticipate there being any significant risks to participants or researchers during this study. It is unlikely that the discussions between participants at the online consensus workshop will involve sensitive topics, as the research tasks are entirely academic in nature. The only likely inconvenience to participants is the time taken to participate in the study. As such, we do not anticipate any undue discomfort or distress to participants during this study. However, should participants express any distress during the online consensus workshop, we will follow the University of Manchester’s Managing Distress policy. This will involve offering the participant the chance to take a break and to leave the workshop without needing to give a reason, and we will emphasise that there are no consequences to withdrawing from the study if participants wish.

## DATA MANAGEMENT PLAN

Data will be collected and stored in accordance with the General Data Protection Regulation (GDPR), the Data Protection Act 2018, the University of Manchester’s [Privacy Notice for Research Participants](https://www.manchester.ac.uk/about/privacy-information/data-protection/privacy-notices/) and Research Data Management Policy. A Data Management Plan for the DeNOTE study is registered with [DMPonline](https://dmponline.dcc.ac.uk/) (ID number 159540).

### Personal information

Participants’ names, place of work, and professional contact details are personal information that is already in the public domain. These details will be collected and stored in a file on the University of Manchester’s secure P:Drive server.

During consent-taking, participants will be given the options of (a) being contacted with the study findings when the research is completed and (b) being contacted about future related research. Both are optional. Once the DeNOTE steering committee have published the research findings and disseminated these findings to any participants who requested a copy, participants’ personal contact details will be destroyed, unless participants consented to be contacted about future related research (in which case, contact details will be securely stored for 5 years and then destroyed).

During consent-taking for the online questionnaire and online consensus workshop, participants will also be given the opportunity to be recognised as a co-author for their role as ‘investigator’ during data construction in the expert workshop. If participants consent to this role, this means that their name and place of work will be personal information that is subsequently reported in the study outputs (e.g., when named as a co-author of a peer reviewed journal article reporting the study and the DN reporting guideline). If participants do not consent to this role, their involvement will be kept anonymous and confidential. However, the identities of participants will necessarily be known to the other participants attending the workshop.

Electronic consent processes (in the online questionnaire and consent form emailed to participants prior to the consensus workshop) will record participants’ names and personal contact details (e.g., email address) so that they can be contacted regarding research activities and study arrangements. The consent files will be stored in a password-protected, encrypted file on the University’s secure P:Drive server for up to 5 years after the research findings have been published, at which point the files will be destroyed. The above is in line with the University’s Research Data Management Policy and Record Retention Schedule.

Only members of the DeNOTE steering committee will have access to the above data. However, individuals from the University of Manchester or regulatory authorities may need to look at the data to make sure that the research is being carried out appropriately.

### Anonymised personal data

Participants’ data generated during the research will be linked to and labelled with an identification (ID) number. Specifically, the ID numbers will be used to anonymise participants’ online questionnaire responses, the online consensus workshop transcript(s), and other data collected during the workshop that can be exported directly (e.g., private voting of the proposed reporting guideline items; any messages sent by participants using the Microsoft Teams ‘chat’ function).

The online consensus workshop will be audio- and video-recorded using Microsoft Teams for later transcription through Microsoft Teams. The transcript will be checked for accuracy using the recordings. All identifying information (e.g., personal, place or organisation names) in the dataset will be replaced or deleted. Participants will therefore be anonymous. The consensus workshop will be recorded and transcribed so that the DeNOTE steering committee have a record of the key topics, ideas, and opinions shared during discussion. This will inform the writing group activities when drafting the DN guideline as well as subsequent study report writing. It is not expected that anonymised participant quotes will be used in study outputs. However, if quotes are used, we will minimise personal identifying information as much as possible to reduce the risk of identifying the participant in the quote. A file containing participants’ names and ID numbers (the ‘key’) will be stored in a password-protected, encrypted file on the University’s secure P:Drive server, separately to all other data collected in the study, which will be stored in password-protected, encrypted files on the University’s Research Data Storage Server.

Should participants wish to withdraw their data, it will be possible to identify their data using their ID number. However, once analysis of the online questionnaire data and, separately, consensus workshop data is completed the files linking participants’ names to their ID number will be destroyed as this data will no longer be needed. Beyond this point, participants will no longer be able to withdraw their questionnaire exercise data or consensus workshop data from the study, as it will no longer be possible to re-identify their data once the file containing their names and linked ID numbers has been destroyed. However, upon request by participants, it may still be possible to redact specific data (e.g., a specific view or opinion shared) after this point, if it can be identified in the consensus workshops transcripts.

Researchers facilitating the discussions at the online consensus workshop will also take field notes. This is to capture the subtle aspects of the interactions and to provide a back-up record of the key topics, ideas, and opinions shared during discussion, in case the audio- and video-recordings fail. These notes will not record any personal identifiable information. This will be checked and any identifiable information will be redacted. It will not be possible to identify and withdraw specific participants’ data from field notes. The field notes will be securely transferred from the facilitators to the lead researcher (HAL) for secure storage and permanently destroyed from the facilitators devices.

The research data will be password-protected, encrypted, and stored on the University’s Research Data Storage Server for up to 5 years after the last research report has been published, at which point the data will be destroyed. Only members of the DeNOTE steering committee will have access to the above data. However, individuals from the University of Manchester or regularly authorities may need to look at the data to make sure that the research is being carried out appropriately.

### Data archive

It is anticipated that the DeNOTE study data will be made publicly available where possible. Participants will be given the option (during consent taking) to have their anonymised data (i.e., online questionnaire responses, online consensus workshop voting responses, consensus workshop transcripts, and ‘chat’ transcript) securely archived in a data repository service such as ReShare (UK Data Service) or Figshare (University of Manchester) for use in future research, teaching, and learning. No personal information will be shared and participants will not be identifiable. This is optional and will only be done with participant’s explicit consent to archive their data. The data of any participants who do not consent to their data being made public will be redacted from the dataset. This redaction will be done after data analysis and before anonymisation (i.e., before the files linking participants’ names to their ID number are destroyed – see **Anonymised personal data** above). *Table 2* indicates the level of data processing, level of access (using terms used by the UK Data Service), and the relevant Rights.

**Table 2.**
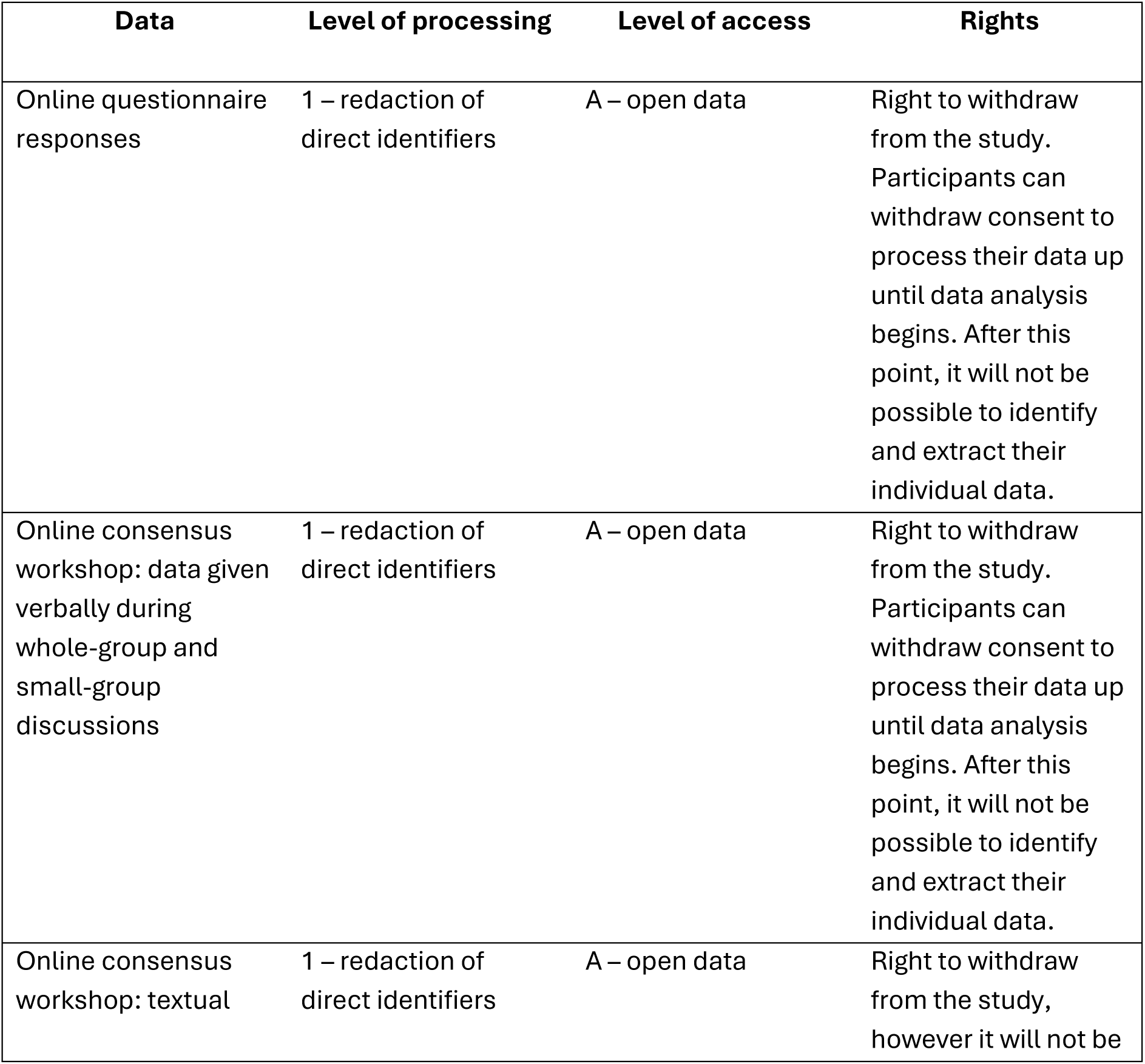
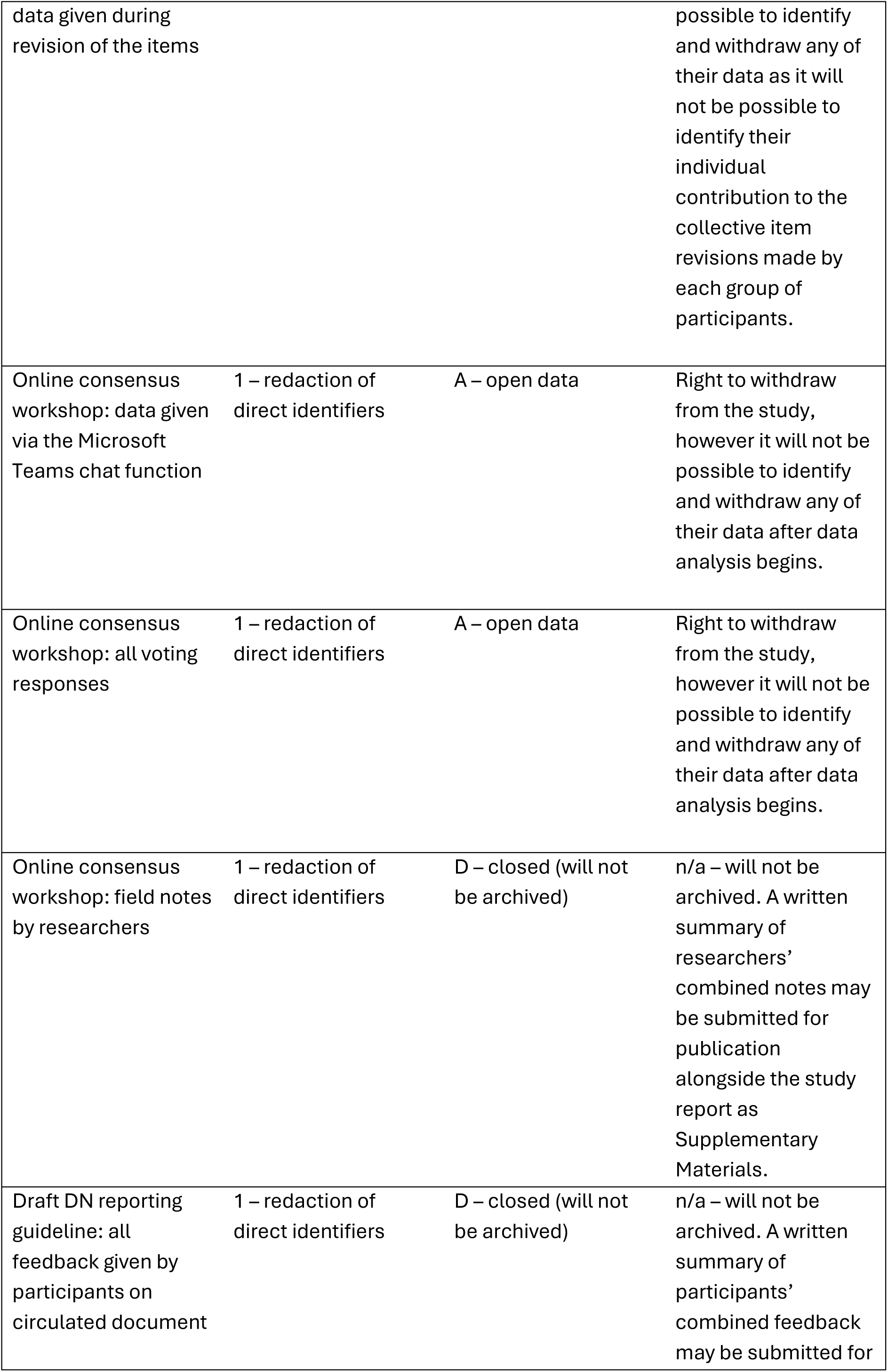
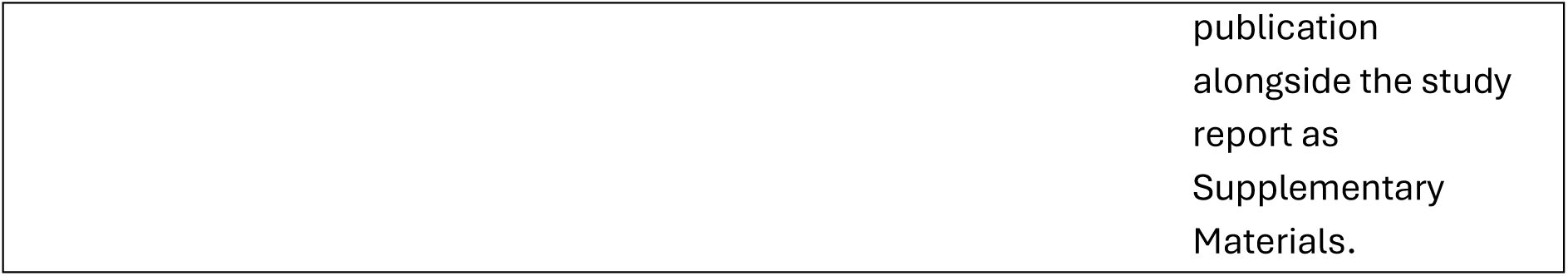
Level of data processing and access.

## DISSEMINATION

The dissemination strategy will be reviewed on an ongoing basis throughout the course of the study to devise the most impactful strategy.

Following study completion, the results will be presented at academic conferences and published in a suitable Open Access peer-reviewed journal. The study article will include the exemplar DeNOTE reporting guideline for uptake and application by others. Study findings may also be disseminated through other mediums and formats (e.g., training materials, blogs). The study registration on the EQUATOR Network will be updated to indicate the reporting guideline has been completed. It is hoped that the DeNOTE reporting guideline will be available via the Network.

It is anticipated that the study data will be made publicly available in a data repository such as ReShare or Figshare. An accompanying DN article describing the archived data will be produced and submitted for Open Access publication in an appropriate journal.

Other dissemination activities are possible. Participants who consent to be classified as an ‘investigator’ for their role in the study will share authorship of all study outputs. As recognised authors, participants can independently disseminate the findings (e.g., at conferences, amongst networks, through teaching activities) and apply for follow-on funding. Thus, this study will form a new expert network who can take further initiative.

## ACKNOWLEDGEMENTS

We are grateful to our Public Contributors (Lynn Laidlaw and Manoj Mistry) who shared their experiences and insights as a public contributor in related research and provided feedback on this study protocol and other study materials.

## DATA AVAILABILITY STATEMENT

As this is a study protocol, we have not collected any data to make available. In time, we intend for our study data to be archived in an appropriate repository. See ‘Data Management Plan’ above.

## APPENDIX A Scopus search string

Searches conducted in Scopus on 11/11/2024

TITLE-ABS-KEY ((“data note” OR “data paper” OR “data article” OR “data descriptor” OR “data report”) AND

(“guide” OR “reporting” OR “template” OR “writing” OR “guidance” OR “guideline” OR “guideline s” OR “framework” OR “frameworks” OR “instruction” OR “instructions”)) AND (LIMIT-TO (LANGUAGE, “English”))

777 documents

141 secondary documents

=918 total.

